# Home-Based Osteoporosis monitoring Using Bioelectrical Impedance Analysis: Muscle-to-Bone Mass Ratio

**DOI:** 10.1101/2023.10.18.23297199

**Authors:** Jingqi Zeng, Xiaobin Jia

## Abstract

Osteoporosis and its associated fractures affect nearly one-fifth of the global population, becoming a significant public health concern. While medical interventions can reduce these risks, Dual-energy X-ray Absorptiometry (DXA), the primary screening method, is limited due to its high cost, need for professional operation, and radiation exposure risks. However, in our Bioelectrical Impedance Analysis (BIA) conducted on 152,449 Chinese residents, we identified two distinct Muscle-to-Bone Mass Ratio (MBR) subgroups in both male and female elderly populations. The MBR range for males is 12.5-15.5 and 16.5-17.8; for females, it’s 11.2-15.2 and 16.5-18.2, with 16 being a significant threshold. Individuals with an MBR value exceeding 16 exhibit pronounced features of osteoporosis and an increased risk of fractures. For instance, the Bone Mineral Content (BMC) in males decreased by 19.18% (95%CI, 18.65%-19.73%), and in females by 29.84% (95%CI, 29.31%-30.36%). These individuals also showed lower body weight and BMI values. Females also displayed shorter stature and a higher body fat percentage, both indicators associated with osteoporosis. In contrast to the traditional Bone Mineral Density (BMD) T-score, the MBR sheds light on a specific physiological state in the elderly. In this condition, their risk of fractures significantly increases. This new understanding of osteoporosis suggests that we should not solely rely on the simple quantification of BMD. Furthermore, BIA measurements taken with domestic scales make MBR a safer, more efficient, and economical screening tool. This enables long-term MBR monitoring at home, offering continuous feedback for osteoporosis and fracture prevention, surpassing the limitations of DXA technology.

## Main

Osteoporosis is one of the most prevalent metabolic bone diseases worldwide, characterized primarily by a reduction in bone mass and alterations in bone microarchitecture. This results in increased bone fragility and an elevated risk of fractures. According to the standards set by the World Health Organization, osteoporosis is defined when the Bone Mineral Density (BMD) at the hip joint or lumbar spine is 2.5 standard deviations (T-score ≤ –2.5) below that of a young adult reference. The global prevalence of osteoporosis stands at 18.35%, with 23.1% in females and 11.7% in males [1]. For individuals aged ≥50 years, a new fracture significantly elevates the risk of subsequent fractures [2], imposing considerable medical and societal burdens. Early pharmaceutical interventions have been demonstrated to effectively alleviate the symptoms of osteoporosis and reduce fracture risks [3,4]. Consequently, emphasizing early screening and risk assessment for osteoporosis holds significant implications for public health.

For self-screening of osteoporosis, one can utilize the one-minute test by the International Osteoporosis Foundation [5] or the osteoporosis self-assessment tool [6]. For high-risk groups, it’s recommended to measure BMD using Dual-energy X-ray Absorptiometry (DXA) to further evaluate the risk of fractures. It’s crucial to recognize that besides a decrease in BMD elevating the fracture risk, other non-BMD factors, such as age, gender, family history, diet, and overall health status, also impact bone health risk [7]. Tools like FRAX®, QFracture, and Garvan integrate these multiple risk factors to predict fracture risk and are essential tools in the clinical management and prevention of osteoporosis [8–10].

BMD is the cornerstone metric for screening osteoporosis and preventing fractures, with DXA being the clinical gold standard. It’s worth noting that skeletal fragility due to endocrine disorders is not always reflected through BMD [11]. In such cases, trabecular bone score [12] and quantitative dual-energy CT [13] can be considered to identify fracture risks. However, the aforementioned BMD and bone microarchitecture testing methods both involve high costs and require skilled professionals, making them challenging to implement widely in resource-limited settings. Moreover, repeated measurements with DXA might raise radiation safety concerns; hence, it’s recommended that the frequency of measurements not exceed once annually. With the increase in average human lifespan, the current osteoporosis screening methods predominantly reliant on DXA might not suffice for public health needs. Consequently, there’s an urgent need to develop safer, more affordable alternatives.

Bioelectrical Impedance Analysis (BIA) is a non-invasive, low-cost, and harmless body composition analysis technique. By analyzing electrical resistance characteristics, it can measure components like muscle, fat, and Bone Mineral Content (BMC) among others and has been widely utilized in assessing nutritional status under health and disease conditions [14,15]. While BMC is influenced by height and weight, making it difficult to establish a T-score system akin to BMD, BIA is still considered the most promising alternative to DXA. The challenge lies in identifying markers from BIA measurements that effectively reflect osteoporosis and fracture risks.

In this study, considering the lack of reference data from DXA, we introduced a novel big data analytic approach that does not rely on the gold standard, termed the Indicator Redirection Strategy (IRS). By analyzing BIA measurements from 152,449 Chinese residents between 2011 and 2017, we identified the Muscle-to-Bone Mass Ratio (MBR) as an effective indicator for assessing osteoporosis and bone health risks in older individuals. MBR can be determined using BIA, offering superior safety, efficiency, and cost-effectiveness. Additionally, given the widespread application of BIA technology in household weighing scales, it provides a convenient and continuous method for long-term home monitoring. This facilitates earlier detection and intervention of latent osteoporosis symptoms, reducing the risk of associated fractures, an advantage not realized by the DXA technique. The overall research design is depicted in **Figure 1**.

**Figure 1.**
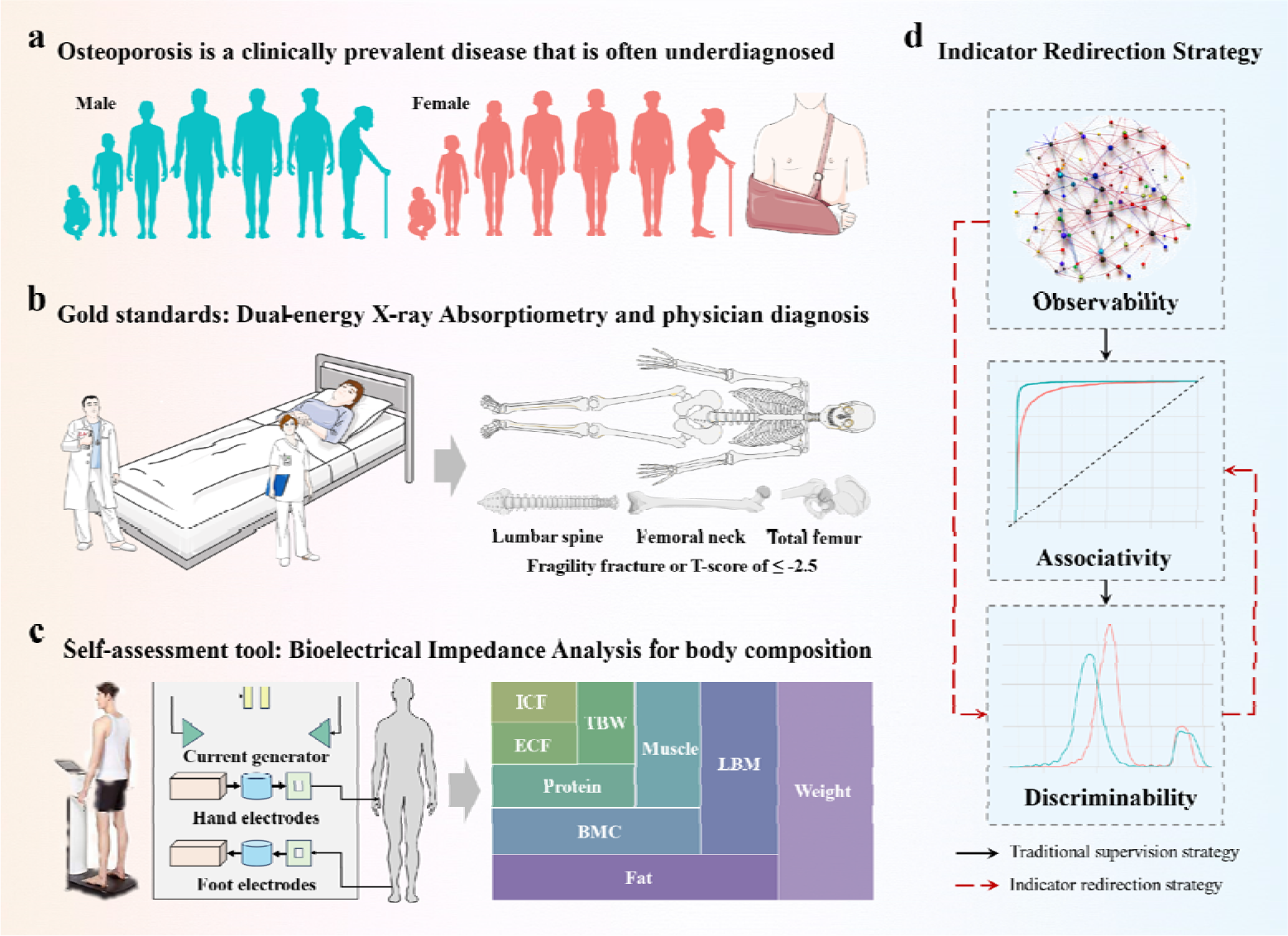
Overall research design. Our aim is to leverage the BIA technology (c) as a substitute for DXA (b) to reduce the cost of osteoporosis screening and fracture risk prediction, enhance safety, and address the growing public health demands (a). As our dataset lacks reference values from DXA, we are unable to employ supervised methods to study indicators associated with BMD (c). Consequently, we introduced the IRS (d). Considering our inability to establish a T-score system analogous to BMD, we expect these indicators to demonstrate a bimodal distribution, reflecting significant characteristic differences within the elderly population. We then further investigate the correlation between these characteristics and osteoporosis as well as fracture risk.

## Results

### Characteristics of Body Composition Data

In this study, we included a total of 152,449 participants, with 80,241 males (52.6% of the total) and 72,208 females (47.4% of the total). The age range of the participants spanned from 6 to 109 years, with those aged 50 and above constituting 18.0% of the total population (Supplementary Figure S1). It’s noteworthy that we encountered missing values in our data for ICF, ECF, and protein. The data completeness for ICF and ECF was both at 79.8%, while for protein it was 88.8%. During data analysis, we opted to exclude the missing values rather than impute or substitute them.

When analyzing gender differences, apart from fat content (males had 0.92 times that of females), all other components were distinctly higher in males, ranging from 1.24 to 1.41 times that of females. For detailed data, please refer to Table 1. Given the significant disparities between genders, subsequent analyses should be conducted separately for males and females.

Of particular interest is that the weight of BMC doesn’t simply decrease with age post-50 (Supplementary Figures S2-S4). In fact, there’s a noticeable dip between the ages of 55 to 70, followed by a swift rise after 70, which then remains stable. This trend is more pronounced among females. Moreover, we discerned that compared to body weight (*r*=0.85), the trend in muscle mass (*r*=0.93) has a stronger correlation with BMC.

**Table 1.** Descriptive Statistics of Human Body Composition and Differences between Genders.

| Variable | N | Min | Max | Mean | SD | Skew | Kurt | Mean<br>(Male) | Mean<br>(Female) | T<br>value | P<br>value |
| --- | --- | --- | --- | --- | --- | --- | --- | --- | --- | --- | --- |
| Age | 152449 | 6 | 109 | 34 | 15 | 0.87 | 0.28 | 33.56 | 34.17 | -7.87 | <0.001 |
| Height (cm) | 152449 | 105 | 220 | 165 | 10 | -0.60 | 1.55 | 171 | 159 | 259.10 | <0.001 |
| Weight (kg) | 152449 | 17.1 | 253.0 | 64.4 | 14.9 | 0.59 | 1.69 | 71.1 | 57.0 | 212.04 | <0.001 |
| Muscle (kg) | 152449 | 10.4 | 173.3 | 45.2 | 10.6 | 0.43 | 0.30 | 52.1 | 37.6 | 368.79 | <0.001 |
| BMC (kg) | 152449 | 0.1 | 10.4 | 3.1 | 0.7 | 0.27 | 0.30 | 3.5 | 2.7 | 305.07 | <0.001 |
| Fat (kg) | 152449 | 0.3 | 79.0 | 16.1 | 6.5 | 0.69 | 1.68 | 15.5 | 16.7 | -35.71 | <0.001 |
| ICF (kg) | 121699 | 3.2 | 89.1 | 24.0 | 5.8 | 0.41 | 0.19 | 27.8 | 19.7 | 346.52 | <0.001 |
| ECF (kg) | 121699 | 2.5 | 46.1 | 11.8 | 2.8 | 0.34 | 0.75 | 13.5 | 9.9 | 295.70 | <0.001 |
| Protein (kg) | 135320 | 2.6 | 38.1 | 10.1 | 2.4 | 0.40 | 0.31 | 11.5 | 8.4 | 338.69 | <0.001 |

### OSTA Screening for Osteoporosis

OSTA is a simple risk assessment tool tailored for the specific characteristics of osteoporosis in Asian populations. It estimates the risk of osteoporosis based on age and body weight. As shown in Figure 2a, there is a direct correlation between age and the rate of high risk as per OSTA, with females always having a higher risk than males. By the age of 85, the risk rate for males is 65.5%, while for females, it reaches 92.5%. This is linked to the fact that females have a lower body weight compared to males of the same age, as depicted in Figure 2b, with their weight being only 80.2% of that of males. Even though the correlation coefficient between BMC and body weight is 0.85, Figure 2c demonstrates that OSTA doesn’t accurately portray the trend of BMC. Especially in females over the age of 50, while their body weight decreases, their BMC remains stable. Assessing the risk of osteoporosis based solely on age and body weight is inadequate. Given the tight connection between BMC and bone mass, our goal is to pinpoint a more precise indicator for osteoporosis and fracture risk based on BMC.

**Figure 2.**
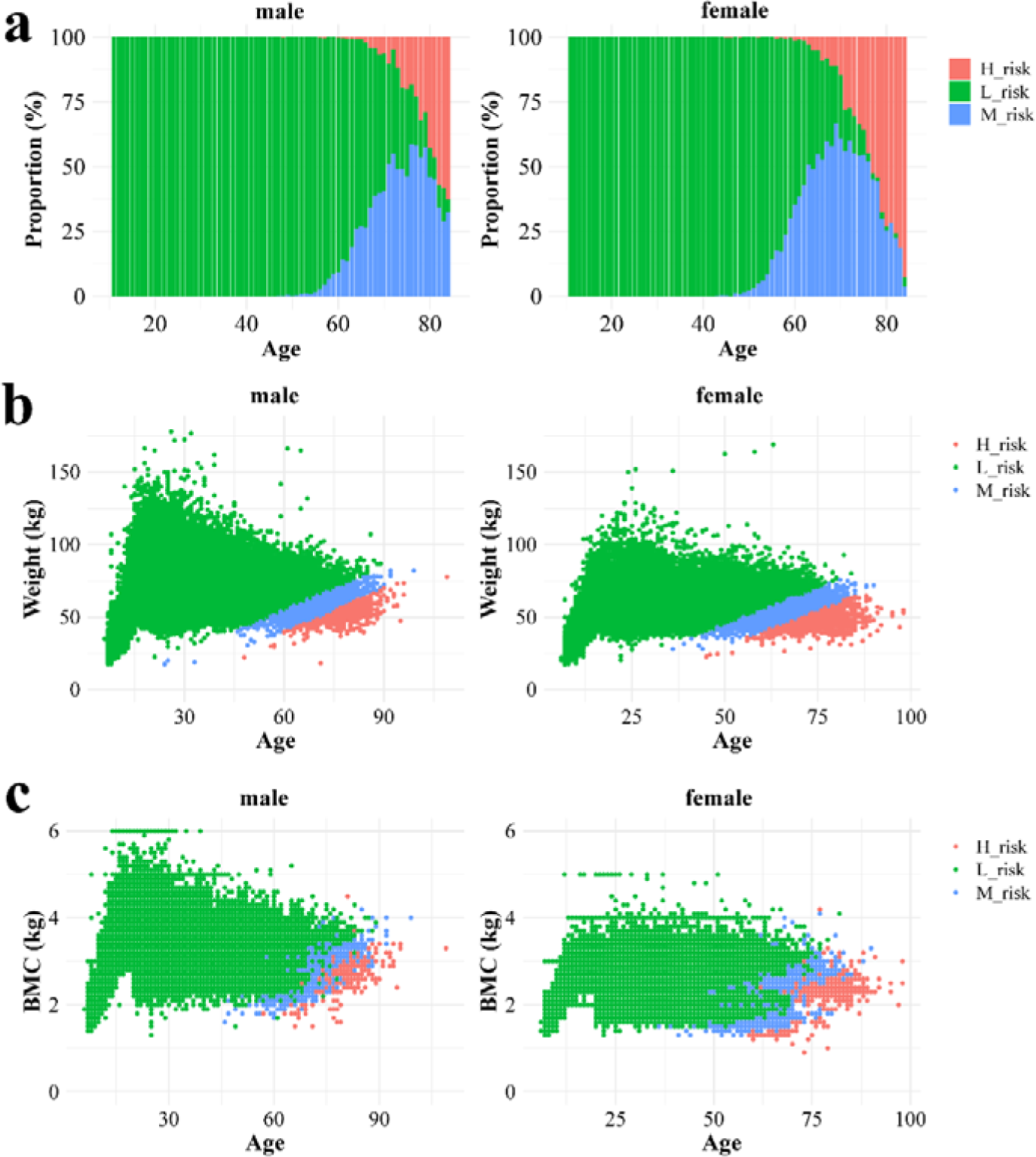
OSTA assessment for osteoporosis. For OSTA values greater than –1, the risk is classified as low; for OSTA values between –1 and –4, the risk is classified as moderate; and for OSTA values less than –4, the risk is deemed high. The percentage of females with high OSTA risk is significantly higher than that of males (a). This can be attributed to females having significantly lower body weights than males within the same age group (b). However, this risk assessment does not entirely capture the characteristics of BMC (c).

### Two Distinct MBR Subgroups Exist in the Elderly Population

Using multi-scale transformation techniques, we derived multiple indicators for BMC and examined their KDE distribution in the elderly population. For instance, when focusing on the weight of BMC (Figure 3a), both men and women exhibit a unimodal distribution, with a range of 1.6-4.5 for males and 1.3-4.2 for females. However, when considering the ratio of BMC to total weight (Figure 3b), females display a bimodal distribution (2.4-4.2 and 3.4-6.5, threshold 3.8), while males maintain a unimodal distribution (3.2-6.2). Inspired by BMI, we introduced a new metric: the ratio of BMC weight to the square of height (Figure 3c). In this metric, the bimodal distribution for females becomes more pronounced (0.6-0.9 and 0.8-1.5, threshold 0.85), while males show a unimodal distribution ranging from 0.7 to 1.5.

**Figure 3.**
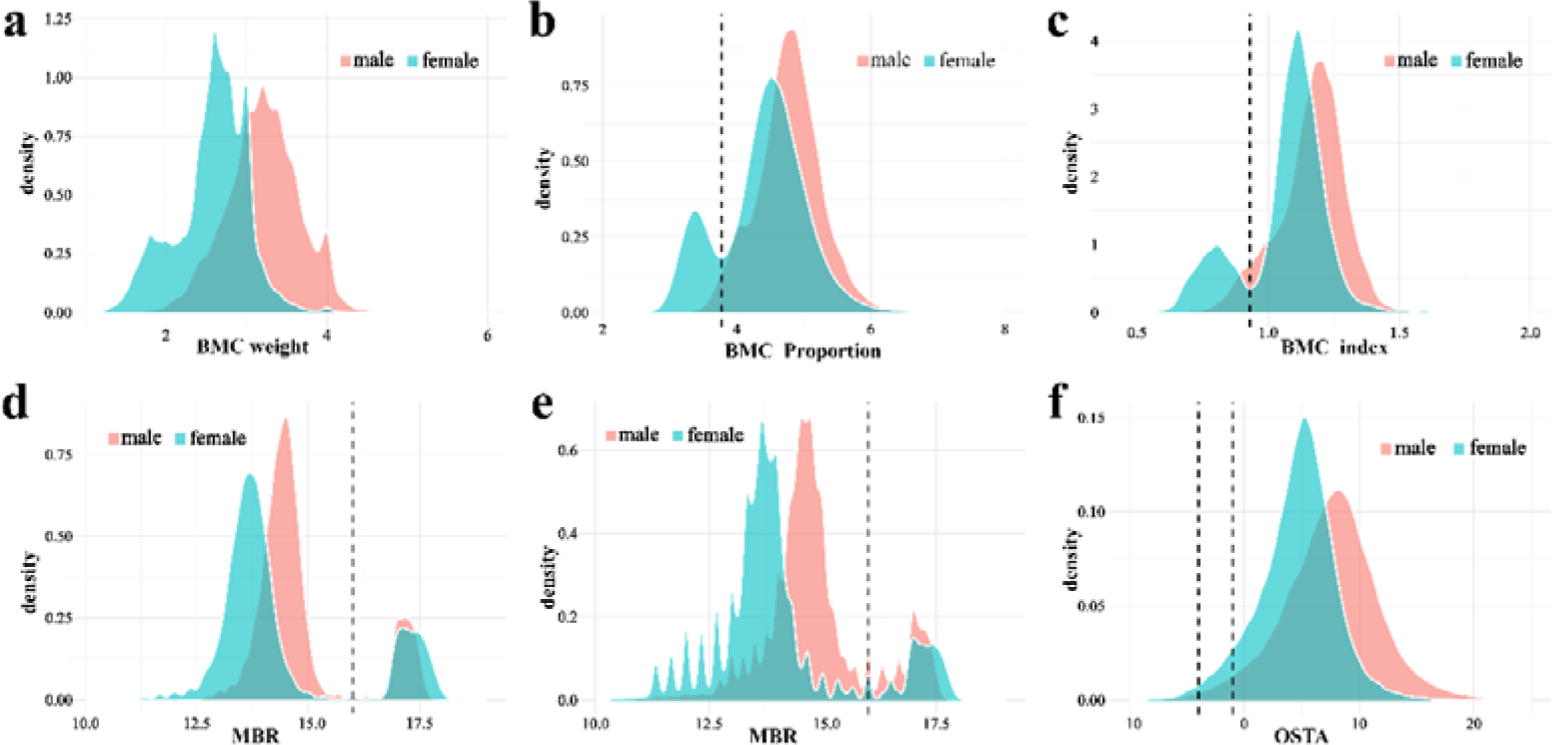
assessment of indicator discriminative power. Within the elderly population, neither the BMC weight (a), the ratio of BMC to total body weight (b), nor the BMC index, which is the ratio of BMC to the square of height (c), exhibited a bimodal distribution for both genders. MBR successfully segregated the population into two distinct subsets by setting a threshold of 16 (d). This distribution pattern is evident across all age groups (e), and its performance is significantly superior to OSTA (f).

Building on our initial observation that muscle trends with BMC consistently over time, we further explored the density distribution of MBR in the elderly. Both males and females exhibit two separate MBR subgroups, as illustrated in Figure 3d. The ranges for males are 12.5-15.5 and 16.5-17.8, while for females they are 11.2-15.2 and 16.5-18.2, with 16 serving as a notable threshold. This bimodal distribution characteristic isn’t confined to the elderly (Figure 3e), but it’s more pronounced in this age group and evidently outperforms OSTA (Figure 3f). We also studied the influence of fat, but found no similar patterns, and its effect on MBR was minimal, as detailed in Supplementary Figures S7-S8. Moving forward, we aim to delve deeper into the compositional differences between these two subgroups and their correlation with osteoporosis and fracture risk.

### Association of MBR with Osteoporosis and Fracture Risk

As shown in Figure 4a, we delved deeper into the differences between the two elderly subgroups segmented by MBR. Notably, individuals with MBR≥16 exhibit characteristics of osteoporosis and are associated with a higher risk of fractures. The most evident data stems from the comparison between BMC and muscle mass. For males in the MBR≥16 group, the BMC weight was reduced by 0.62-0.65 kg, a decrease of 19.18% (95%CI, 18.65%-19.73%); while for females, it was reduced by 0.80-0.83 kg, a drop of 29.84% (95%CI, 29.31%-30.36%). Concurrently, muscle mass in males decreased by 1.53-2.12 kg, a 3.81% reduction (95%CI, 3.19%-4.43%); and in females, it was down by 4.00-4.48 kg, an 11.30% reduction (95%CI, 10.67%-11.94%). This trend is supported by data from ratio and index transformations. Furthermore, individuals in the MBR≥16 group were younger on average: males were 1.61 years younger (95%CI, 1.30-1.92) and females were 1.79 years younger (95%CI, 1.53-2.06), eliminating the impact of age.

**Figure 4.**
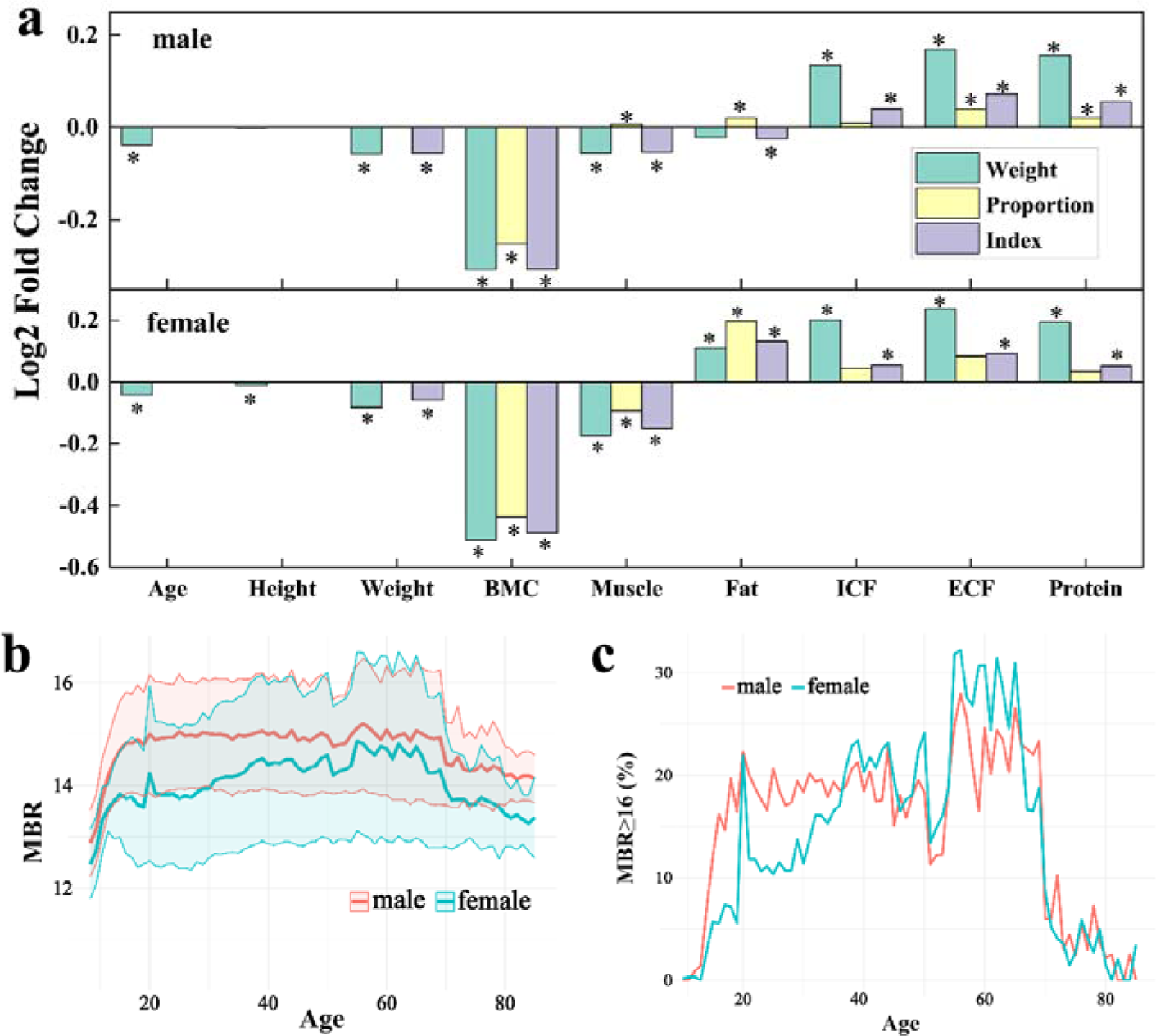
MBR Assessment for Osteoporosis. (a) A comparison was made between the elderly population with an MBR≥16 and other groups in terms of various indicators, including weight, proportion, and data after index conversion. In the figures, the vertical axis represents the logarithmically transformed values in terms of fold change, while “*” indicates statistically significant differences. (b) The trend of MBR changes with age. (c) The proportion of the population with MBR≥16 changes with age.

Additionally, the MBR≥16 group displayed osteoporosis-related features. Weight was reduced by 2.13-3.18 kg in males, a 3.89% decrease (95%CI, 3.12%-4.66%), and by 2.90-3.76 kg in females, a 5.56% reduction (95%CI, 4.84%-6.28%). There was also a drop in BMI: 0.78-1.10 in males, a 3.79% decrease (95%CI, 3.13%-4.64%), and 0.83-1.14 in females, a 4.00% drop (95%CI, 3.38%-4.82%). Females also showed a decrease in height of 1.10-1.62 cm, a 0.87% drop (95%CI, 0.71%-1.05%). Additionally, fat increased by 1.34-1.81 kg in females, an 8.00% increase (95%CI, 6.81%-10.67%), possibly related to hormonal imbalances post-menopause. We also observed increases in ICF, ECF, and protein, but their proportions didn’t exhibit significant changes, which may correlate with chronic inflammation and physiological adjustments (Supplementary Figure S9).

While MBR is derived from both muscle and BMC metrics, it primarily reflects changes in BMC rather than muscle (Supplementary Figure S10). Specifically, the AUC values for BMC weight, ratio, and index in males were 0.89, 0.95, and 0.95 respectively, and for females, they were 0.98, 0.99, and 0.99. In contrast, the association between MBR and muscle metrics was weaker. AUC values for muscle weight, ratio, and index in males were 0.59, 0.51, and 0.62, and for females, they were 0.75, 0.72, and 0.78.

Analyzing the change of MBR with age, the age range of 50-55 years emerges as a crucial window for osteoporosis intervention. Within this span, the average MBR sees a rapid decline, then peaks at the age of 55, and undergoes a swift decay post the age of 70 (as visualized in Figure 4b). This trajectory is evident in the proportions of the MBR≥16 group: as shown in Figure 4c, between 50-55 years, the proportions for males and females stand at 13.5% and 17.7% respectively. This climbs to 23.5% for males and 28.8% for females in the 55-70 age bracket, but drops sharply to 5.2% for males and 4.3% for females between 70-85 years. This pattern aligns well with the age-related changes in BMC.

We hypothesize that, upon reaching the age bracket of 50-55 years, the BMC starts to decrease due to bodily aging. However, internal stress feedback mechanisms temporarily stimulate an increase in BMC, subsequently influencing the MBR value. Consequently, clinical interventions targeting this specific age range could potentially offer effective osteoporosis prevention and reduce fracture risks. It’s noteworthy, however, that for individuals over 70 years, MBR’s applicability has its limitations. For this age group, judgments might rely more heavily on BMD readings.

In summary, MBR, as a comprehensive indicator, unveils a specific physiological state in the elderly. In this state, there is a significant correlation between BMC and muscle mass, and it is also characterized by low body weight, low BMI, shorter stature in females, and a high body fat percentage—all indicative of osteoporosis. With this condition, their risk of fractures significantly increases. This newfound understanding of osteoporosis suggests that we should not solely rely on the simple quantification of BMD.

## Discussion

In our study, based on the bioelectrical impedance analysis (BIA) measurements of 152,449 Chinese residents, we investigated the age-related changes in body components such as muscle, BMC, and fat. We observed a bimodal distribution pattern in the MBR within the elderly population, which distinctly outperforms the currently utilized self-assessment tool, OSTA. The subgroup with MBR≥ 16 exhibited characteristics of low BMC and muscle mass, further establishing MBR as an innovative and effective marker for evaluating the risks of osteoporosis and fractures.

The Index of Risk Score (IRS) represents an avant-garde paradigm in medical risk assessment, propelled by the burgeoning realm of big data [16–17]. Nevertheless, the dearth of a definitive clinical benchmark curtails traditional supervised models. We contend that a salient risk evaluation index should encapsulate three tenets: Observability (ensuring intuitive brevity), Discriminability (adept distinction between varied populations or states), and Associativity (retaining its biological or clinical pertinence). The IRS methodology underscores the discriminative acumen of an index while probing its disease or condition correlations, fostering the genesis of germane metrics. Notwithstanding validation hurdles, IRS’s ingenuity, particularly amidst data paucity, augments the scholarly landscape of medical research.

MBR, considering the combined effects of muscle and BMC, reflects a specific physiological state in the elderly, providing us with a fresh perspective on osteoporosis. In the elderly, osteoporosis and sarcopenia are two prevalent coexisting health issues. These two problems share various risk factors, such as genetics and endocrine imbalances [18]. Biologically, an intricate nexus intertwines muscles and bones. Specific myogenic mediators, exemplified by IL6 and IGF-1, are instrumental not only in osseous vitality [19] but also intimately affiliate with osteocyte functionalities [20]. Given this symbiosis, prevailing BMD-centric evaluative paradigms often marginalize muscle mass’s import. The MBR≥ 16 demographic, delineated by conspicuous diminutions in BMC and muscle mass, emerges as a vanguard stratified for osteoporosis detection and fracture risk prognostication.

The population group with MBR≥16 exhibited other traits associated with osteoporosis, such as low body weight, decreased BMI, reduced height especially in women, and increased fat – all of which are recognized markers of osteoporosis risk [8–10]. Some self-assessment tools gauge the risk of osteoporosis based on age and weight [21–23]. A reduction in height, particularly due to vertebral compression fractures, is regarded as an indicator of osteoporosis risk [24]. The increase in fat content is linked to postmenopausal hormonal changes. Estrogen can stimulate mesenchymal stem cells to differentiate towards osteoblasts, but in osteoporotic patients, this differentiation tends to favor adipocytes [25, 26]. Furthermore, high fat content may elevate the risk of conditions like metabolic syndrome, cardiovascular diseases, and diabetes [27]. This underscores that MBR is not only associated with osteoporosis and fracture risks but may also reflect other health risks postmenopausal women face due to hormonal imbalances.

Osteoporosis, delineated by a BMD T-score ≤-2.5, inherently aims to prognosticate fracture susceptibility. Within this ambit, our MBR criterion potentially stands superior in presaging fracture propensities. MBR screenings underscore an osteoporosis prevalence of 23.5% in males and 28.8% in females within the 55-70 age bracket. Conversely, extant Chinese epidemiological insights via DXA scanning elucidate [28] a 5.4% and 37.1% osteoporosis incidence in 60-70-year-old males and females, respectively, with vertebral fracture prevalence registering at 14.3% and 15.5%. This pronounced variance intimates that MBR, rather than merely echoing bone density metrics, may more acutely mirror genuine fracture susceptibilities. Notably, in males, MBR’s predictive acumen resonates more congruently with actual vertebral fracture statistics. Thus, while DXA retains its primacy for osteoporosis diagnosis, MBR might proffer a refined, pragmatic instrument to gauge authentic fracture peril.

MBR conspicuously outperforms BMD in osteoporosis surveillance and peril assessment. Foremost, MBR’s reliance on the BIA paradigm, vis-à-vis DXA, enhances safety, expediency, and fiscal prudence in clinical contexts. This ascendancy is palpable in expansive osteoporosis epidemiological ventures, heralding a pragmatic modus operandi. Furthermore, the incorporation of BIA into domestic weighing apparatuses bequeaths seniors an effortless self-evaluation conduit, facilitating uninterrupted longitudinal oversight. This paradigm is poised to preemptively discern and counter osteoporosis’ insidious manifestations, mitigating concomitant fracture vulnerabilities — a realm elusive to DXA methodologies. Although a plethora of research endeavors [29–32] have ventured into BIA’s applicability for osteoporosis appraisal, encompassing metrics like body cell mass and phase angle, MBR’s evaluative prowess remains unparalleled.

Using MBR, we provide a more precise method for elderly osteoporosis management and fracture risk assessment, offering patients a comprehensive intervention strategy. Proposed applications include: Mass Screening: Employ MBR and BIA for early fracture risk assessment in the high-risk group aged 55-70. Personalized Management: With household BIA devices, patients can self-evaluate MBR, achieving long-term osteoporosis monitoring. Early Intervention: Based on MBR feedback, doctors can promptly develop and optimize intervention measures to reduce fracture risk. Public Education: Centered on MBR, strengthen public health education for osteoporosis and fracture prevention.

However, our research has limitations: Geographical Scope: It’s primarily based on the Chinese population and needs validation in other regions. Nature of the Study: It’s currently cross-sectional; further exploration is required to understand the long-term association between MBR, osteoporosis, and fractures. Data Completeness: To ensure the widespread application of MBR, more comprehensive data support is required.

In conclusion, our findings preliminarily prove the value of MBR in screening for osteoporosis and assessing fracture risk, but further research is necessary to ensure its broader application.

## Methods

### Body Composition Dataset of the Chinese Population

The body composition dataset of the Chinese population is sourced from the “National Population Health Science Data Center’s Data Repository PHDA.” Between 2011 and 2017, this dataset compiled body composition data from 152,449 Chinese individuals, encompassing muscle, BMC, fat, Intracellular Fluid (ICF), Extracellular Fluid (ECF), and protein, among others. To ensure data accuracy, we compared isotopic labeling results from 32 healthy adults with BIA outcomes. The correlation for TBW was 0.962 (H2^18O) and 0.941 (D2O). Concurrently, the difference between the Magnetic Resonance Imaging (MRI) measurements of fat and skeletal muscle from 140 samples and BIA was 0.89%. In comparison, the difference between the DXA measurements of bone mineral content from 669 samples and BIA was 0.98%. These results validate the reliability of BIA measurements.

### Osteoporosis Self-assessment Tool for Asians

The Osteoporosis Self-assessment Tool for Asians (OSTA) primarily evaluates the risk of osteoporosis based on age and body mass. OSTA calculation method is derived by subtracting the product of age in years multiplied by 0.2 from body weight in kilograms. Based on the OSTA score: OSTA > –1 indicates a low risk. –1 > OSTA > –4 indicates a moderate risk. OSTA < –4 indicates a high risk.

### Kernel Density Estimation

Kernel Density Estimation (KDE) is a non-parametric method used to estimate the probability density function of a continuous random variable. The principle of this method relies on the local characteristics of data points and utilizes a kernel (often the Gaussian kernel) to smooth the data, resulting in a continuous probability density estimate. Mathematically, KDE can be represented as:

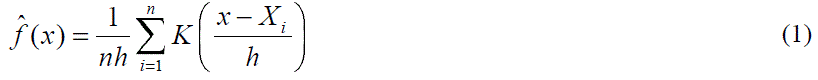

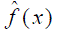 is the density estimation at point x. *K* represents the kernel function.

*h* is the bandwidth determining the degree of smoothness.

*X_i_* denotes individual data points. *n* stands for the number of data points.

## Statistical Analysis

All analyses were conducted in R software (version 4.2.2). Data visualization was achieved using ggtern (version 3.4.2) and ggplot2 (version 3.4.3). In two-sided statistical tests, a P-value of <0.05 was considered to be statistically significant.

## Data availability

The human body composition dataset for the Chinese population can be accessed through the National Population Health Data Center (https://www.ncmi.cn/) under the Creative Commons – Attribution 4.0 International license. The data can be accessed directly via the following link: https://www.ncmi.cn//phda/dataDetails.do?id=CSTR:A0006.11.A0005.201905.000346.

## Author contribution statement

**Jingqi Zeng:** Experiment, Data curation, Formal analysis, Investigation, Methodology, Software, Validation, Visualization, Writing – original draft, Writing – review &editing. **Xiaobin Jia:** Conceptualization, Funding acquisition, Project administration, Supervision, Writing-review &editing.

## Competing interests

The authors declare that they have no known competing financial interests or personal relationships that could influence the work reported in this paper.

## Acknowledgements

The body composition dataset of the Chinese population was sourced from the’ National Population Health Data Center’. This research was supported by the National Natural Science Foundation of China (Grant No. 82230117,China). The funders of the study had no role in study design, data collection, data analysis, data interpretation or writing of the report.

## Supplementary information

We present figures detailing the dataset’s characteristics (Figure S1) and age-related changes in body composition (Figures S2-S4). The elderly demographic and its various indicators are examined (Figure S5), alongside discriminative capacities for indicators, especially BMC (Figures S6-S7). Proportional associations of key body components are covered in Figures S8 and S9. Lastly, Figure S10 showcases the ROC curves for BMC and muscle in relation to MBR.

## Notes

### Competing Interest Statement

The authors have declared no competing interest.

### Author Declarations

The human body composition dataset for the Chinese population can be accessed through the National Population Health Data Center (https://www.ncmi.cn/) under the Creative Commons - Attribution 4.0 International license. The data can be accessed directly via the following link: https://www.ncmi.cn//phda/dataDetails.do?id=CSTR:A0006.11.A0005.201905.000346.

### Summary of Updates

We have made revisions to our manuscript's abstract and certain sections of the main text to enhance clarity and accuracy. Please be assured that the modifications made to our manuscript are consistent with the core findings and conclusions of our original submission.

