## Supplementary information for "Home-Based Osteoporosis monitoring Using Bioelectrical Impedance Analysis: Muscle-to-Bone Mass Ratio" (from another version of this preprint: version 1, 2023-10-19)

### Figure S1. Basic characteristics of the dataset

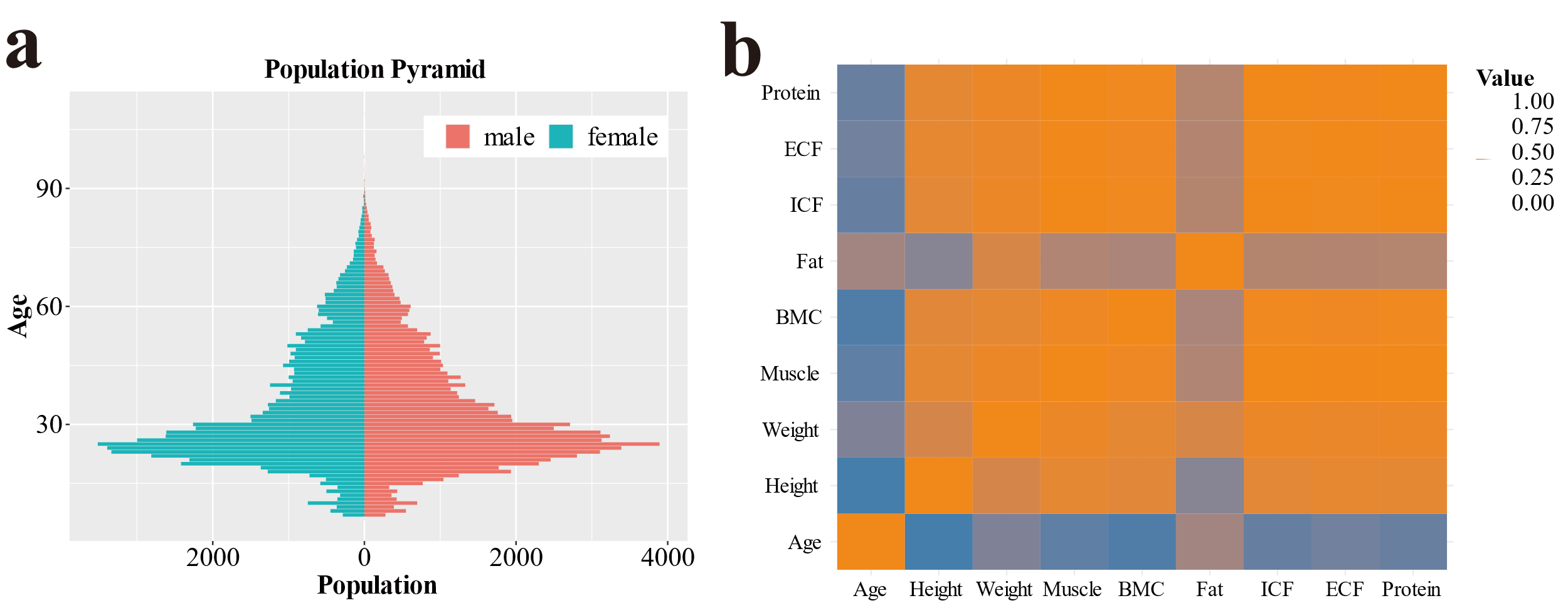

(a)Demographics of survey participants. (b)Pearson correlation coefficients of various components. An extensive assessment of body composition was conducted on 152,449 Chinese citizens, comprising 80,241 males (52.6%) and 72,208 females (47.4%). Of these, 27,327 individuals were aged 50 and above, making up 18.0% of the sample. Except for fat, all components exhibited strong positive correlations.

### Figure S2. Changes in body composition weight with age

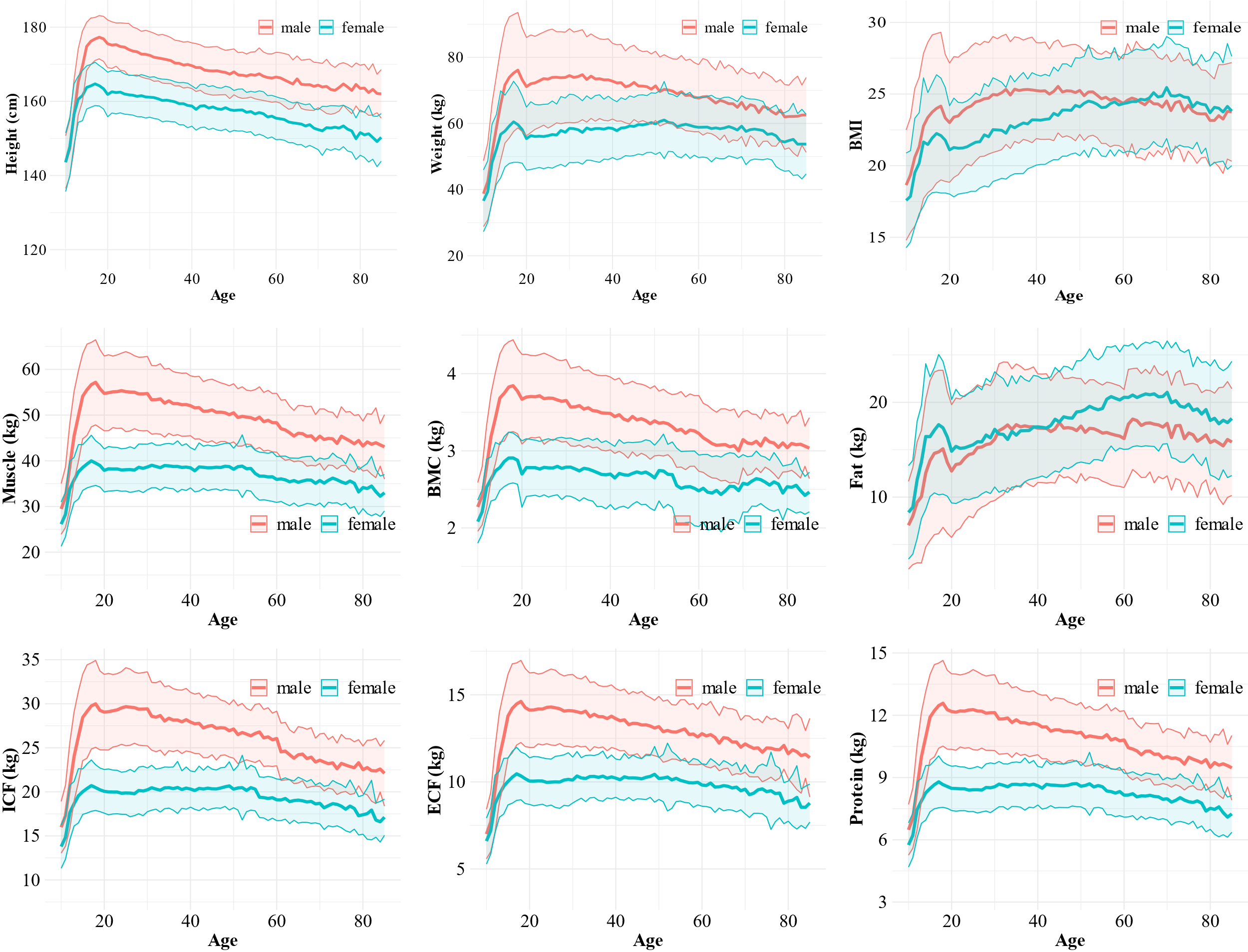

The dataset includes indicators such as height, weight, BMI, muscle mass, BMC (Bone Mineral Content), fat, ICF (Intracellular Fluid), ECF (Extracellular Fluid), and protein. Apart from BMI and fat, men's other indicators in various age groups are higher than those of women. The age range of 55-70 years is a crucial period when significant changes in body components such as muscle, BMC, and fat are observed.

### Figure S3. Changes in body composition proportion with age

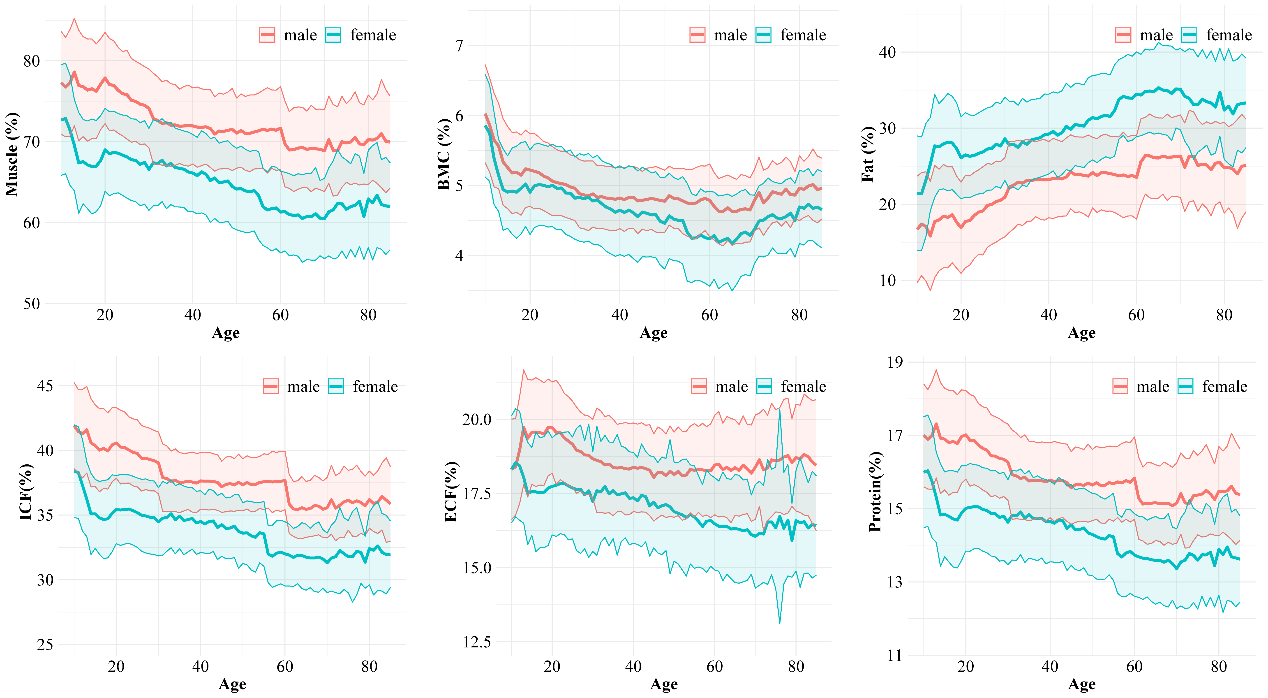

The dataset includes body components such as muscle, BMC, fat, ICF, ECF, and protein. The data displays the proportion of each body component in relation to the total body weight. The age range of 55-70 years is a critical period for changes in body composition.

### Figure S4. Changes in body composition index with age

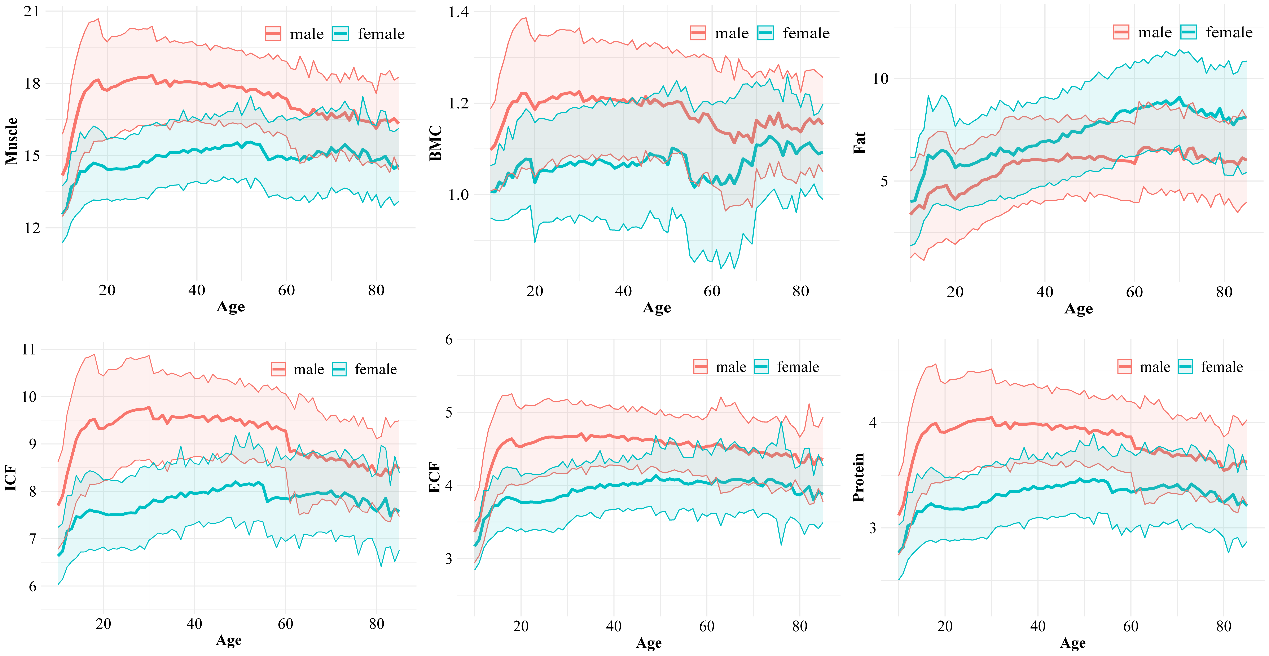

The dataset includes body components such as muscle, BMC, fat, ICF, ECF, and protein. The data shows the ratio of the weight of each body component to the square of height. During the age range of 55-70 years, significant changes in body composition are also observed.

### Figure S5. Density distribution of various indicators in elderly population

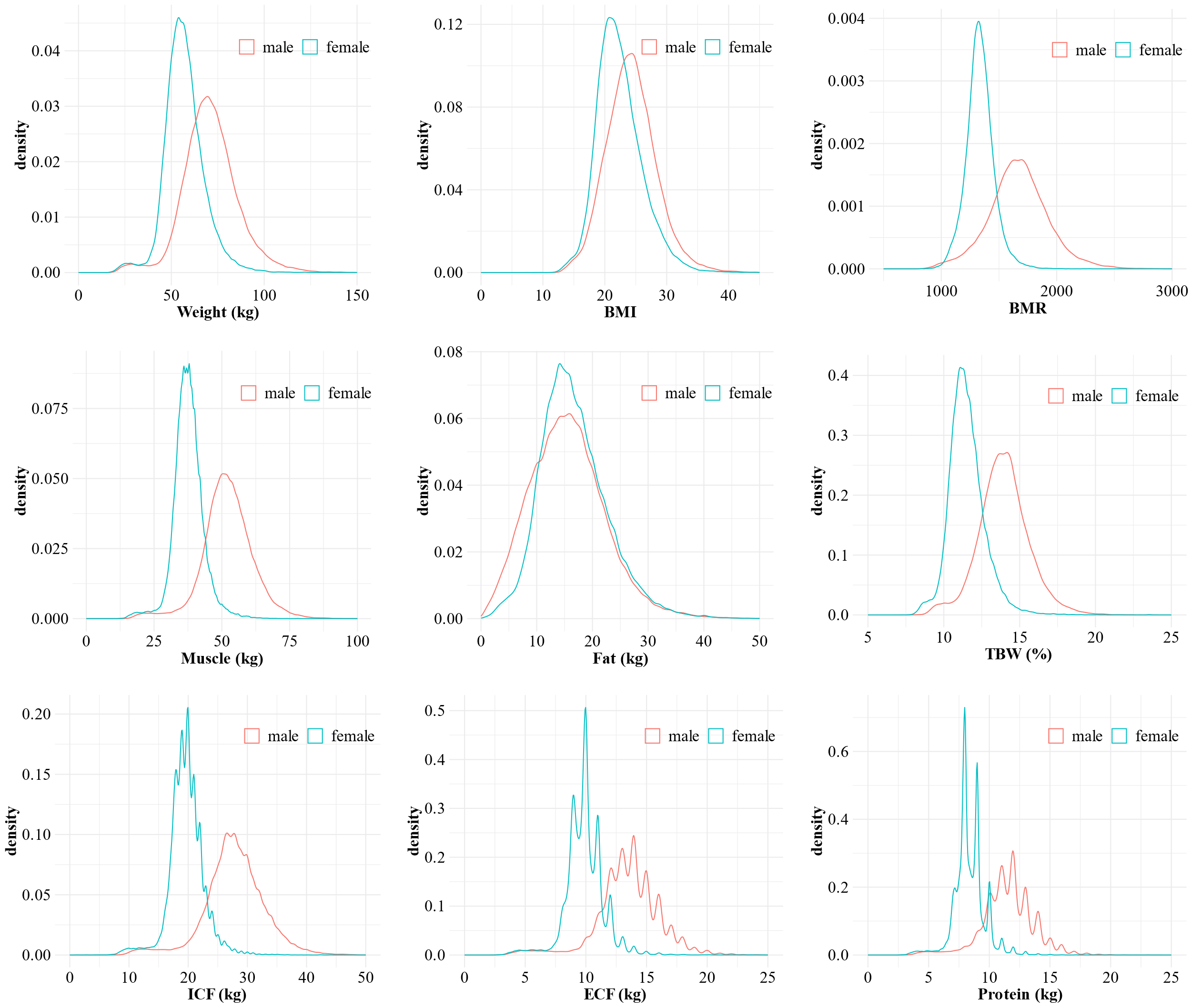

Density distribution of several body composition indicators in the elderly population (those aged over 50 years). The indicators primarily show a unimodal distribution or an unstable multimodal distribution. However, the distinction between peaks is minimal, insufficient for significant population differentiation.

### Figure S6. Discriminative capacity of the indicator for BMC across all age groups.

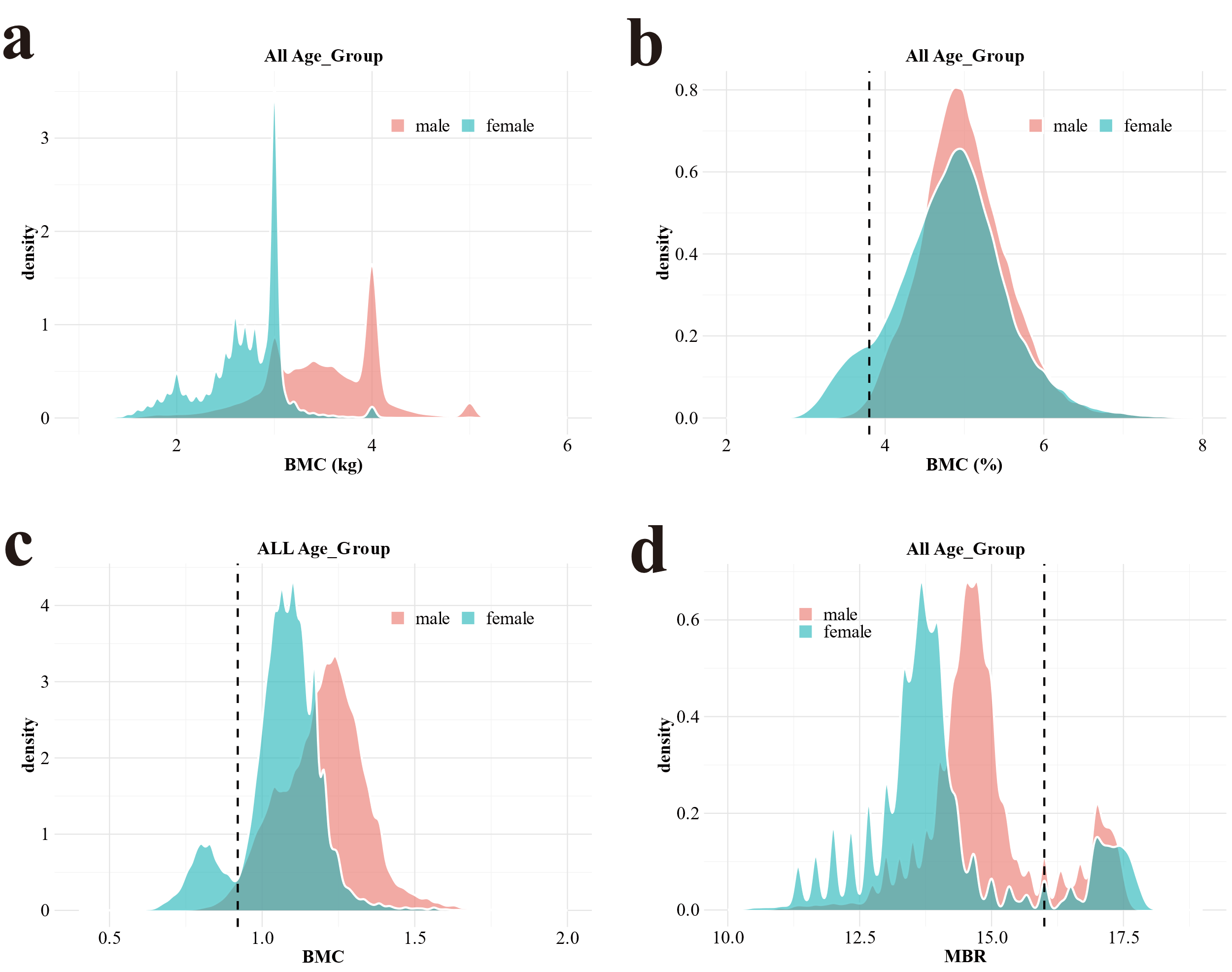

(a) Distribution of BMC weight, showing limited differentiation in the elderly population. (b) Distribution of the proportion of BMC in total weight. There's a bimodal pattern, yet it does not meet our expectations. (c) Distribution of BMC data normalized by height squared. It offers good differentiation in females but is less effective for elderly males. (d) Distribution characteristics of MBR, presenting a distinct bimodal distribution in both males and females.

### Figure S7. Discriminative capacity of the indicator for the weight ratio of fat to BMC

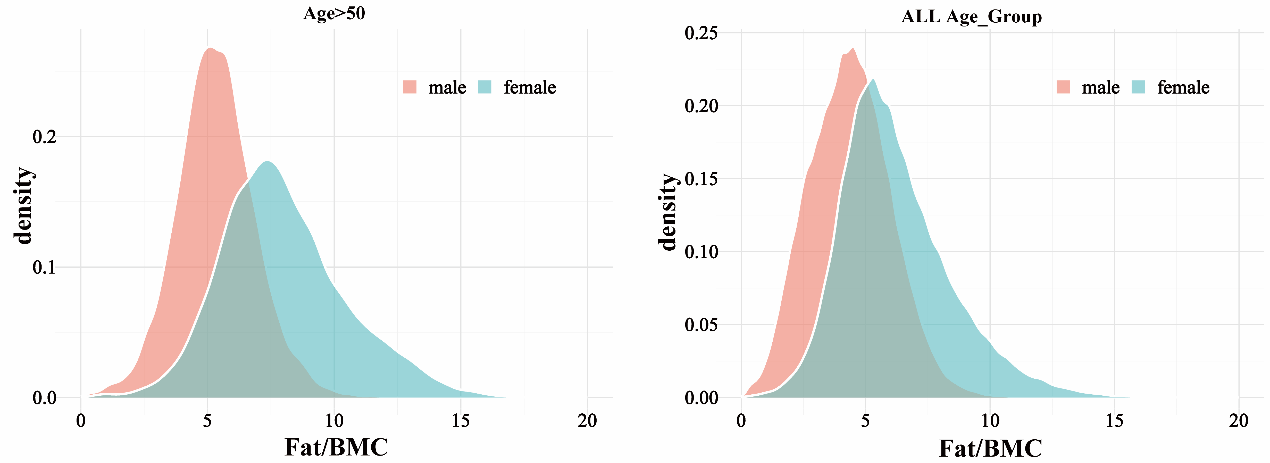

The ability of the weight ratio of fat to BMC to differentiate between the elderly population and the overall age group was evaluated. Clearly, it does not possess characteristics similar to MBR, and cannot accurately segment the population.

### Figure S8. Proportional association of muscle, BMC, and fat in the human physique

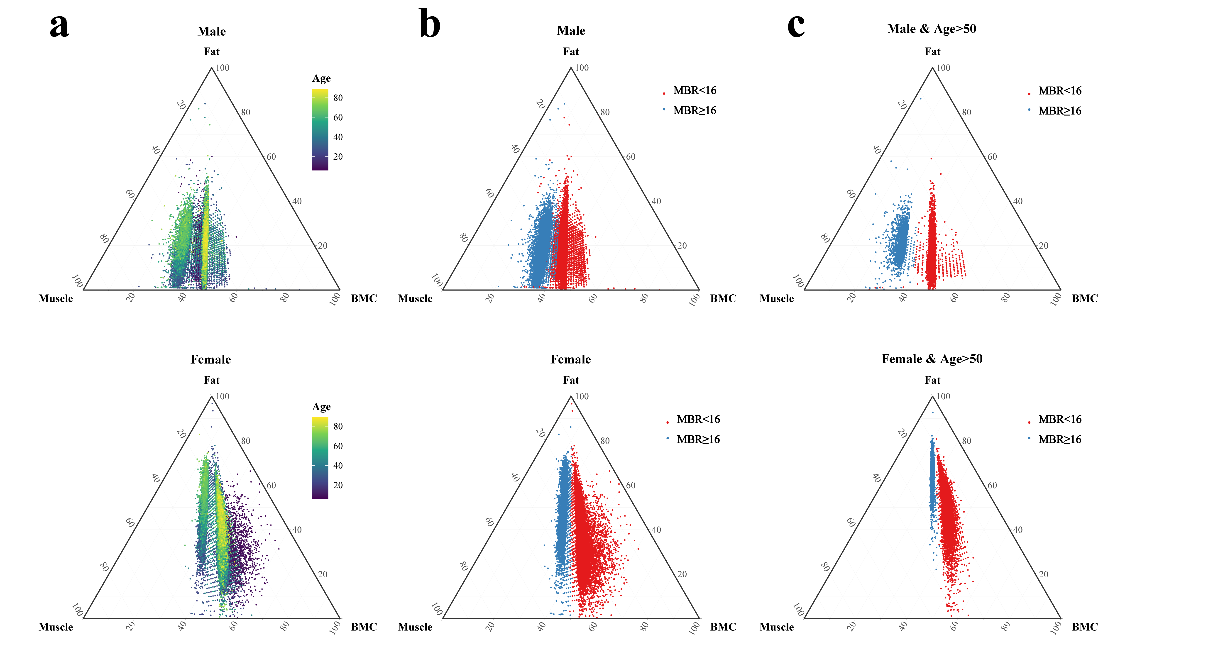

(a) For all age groups, the coloring of the dots is based on age. The coloring of the dots is based on MBR for all age groups (b) and the elderly population (c). Emphasizes the discriminative effect when MBR≥16. Fat has a minor influence on the proportion of muscle to BMC. MBR≥16 exhibits significant differentiation abilities in both males and females.

### Figure S9. Proportional association of ECF, ICF, and protein in the human physique

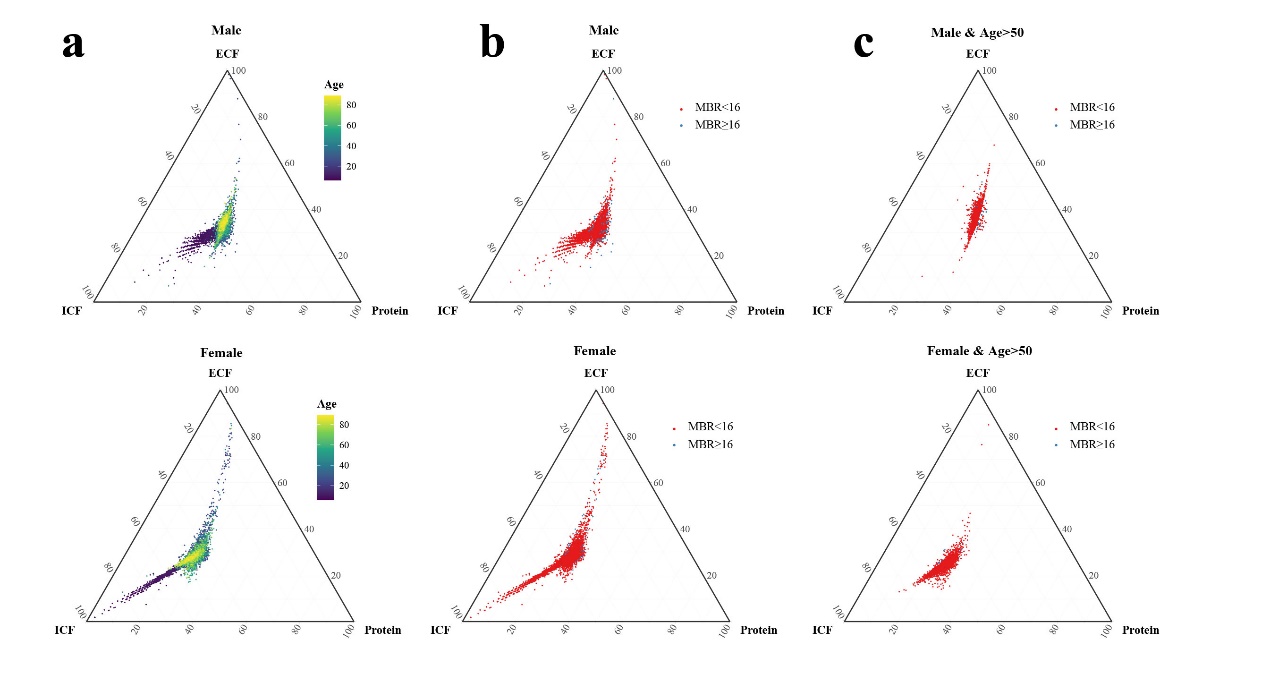

(a) For all age groups, the coloring of the dots is based on age. The coloring of the dots is based on MBR for all age groups (b) and the elderly population (c).Dividing populations based on MBR≥16, the proportions of these three components show no significant difference among different groups.

### Figure S10. ROC curves for BMC and muscle in relation to MBR.

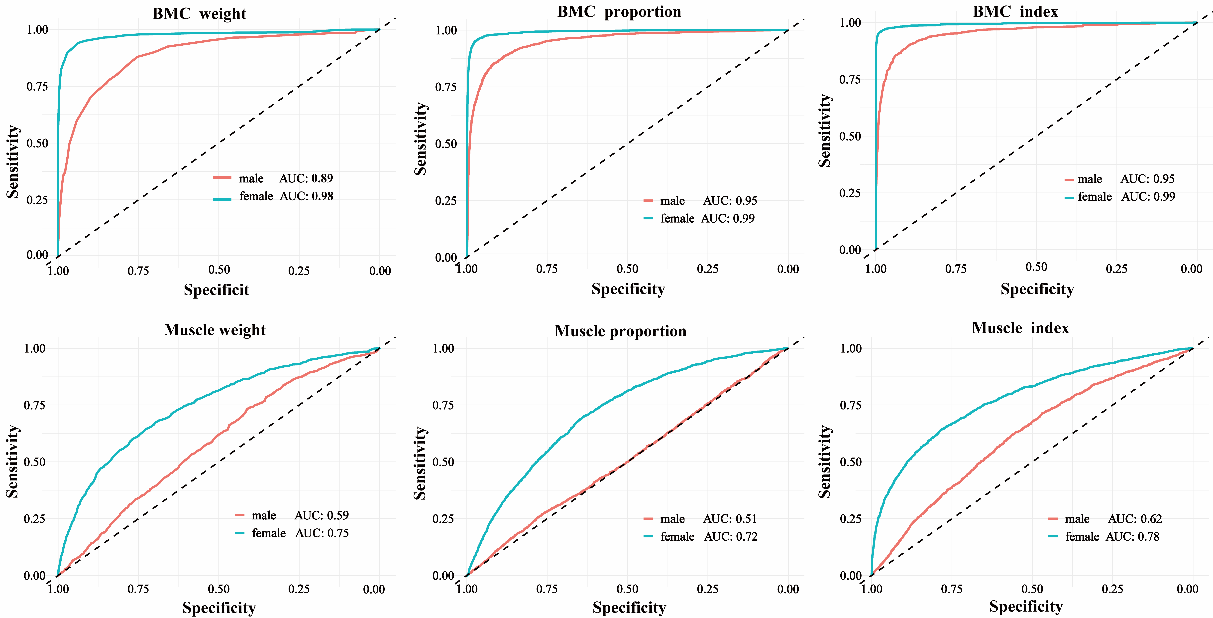

The AUC values for BMC weight, proportion, and index in males are 0.89, 0.95, and 0.95 respectively, while for females, they are 0.98, 0.99, and 0.99. In contrast, the association between MBR and muscle indicators is weaker. The AUC values for muscle weight, ratio, and index in males are 0.59, 0.51, and 0.62, whereas for females, they are 0.75, 0.72, and 0.78.
